# Onca: An Open 9B Language Model for Pancreatic Cancer Clinical Tasks

**DOI:** 10.64898/2026.04.16.26351055

**Authors:** Kwan Bo Shim

## Abstract

Pancreatic ductal adenocarcinoma (PDAC) remains one of the deadliest solid tumors and continues to face low treatment-trial participation, fragmented evidence workflows, and labor-intensive abstraction of unstructured clinical text. Existing oncology-focused language models show promise, but many depend on private institutional corpora, limiting reproducibility and practical reuse across centers. We present **Onca**, an open 9B dense model designed for four PDAC-relevant tasks: trial eligibility screening, case-specific clinical reasoning, structured pathology report extraction, and molecular variant evidence reasoning. Onca is fine-tuned from **Qwopus3.5-9B-v3** with a single Unsloth BF16 LoRA adapter on 37,364 training rows drawn from openly available sources. The evaluation spans 11 panels and compares Onca against Woollie-7B, CancerLLM-7B, OpenBioLLM-8B, and the unmodified Qwopus base. Onca achieves the strongest overall results on Trial Screening (81.6 F1), Clinical Reasoning (14.1 composite), Pathology Extraction (30.5 field exact-match), PubMedQA Cancer (68.3 macro-F1), and PubMedQA (66.5 macro-F1). The strongest gains appear in tasks closest to routine oncology workflow, especially trial review and pathology structuring. These findings suggest that clinically targeted pancreatic-cancer language models can be built from open data with competitive performance while remaining practical to train on a single workstation-scale GPU setup.

## 1. Introduction

Pancreatic ductal adenocarcinoma (PDAC) is now the third leading cause of cancer death in the United States, projected to overtake colorectal cancer as the second by the end of the decade [Siegel et al., 2023]. The five-year survival rate remains approximately 13%, the lowest of any major solid tumor, and more than 80% of patients present with locally advanced or metastatic disease at diagnosis [Mannucci et al., 2026]. Improving outcomes therefore depends not only on new therapies but also on operational improvements across the clinical workflow: faster identification of patients who could benefit from clinical trials, more consistent treatment reasoning that incorporates rapidly evolving evidence, and more reliable structured abstraction of the diagnostic information that drives downstream care.

Large language models are a natural candidate for these workflows, but the current oncology-LLM literature has a consistent reproducibility gap. Woollie [Li et al., 2025a,b] reports strong cancer-progression prediction (AUROC 0.97) using 39,319 radiology impression notes from 4,002 Memorial Sloan Kettering patients, but its training corpus is institutional and cannot be redistributed. CancerLLM [Li et al., 2026] reports F1 91.78% on cancer phenotype extraction using the University of Minnesota Clinical Data Repository, comprising 2,676,642 clinical notes and 515,524 pathology reports across 17 cancers, again without public release of the training data. OpenBioLLM-8B [Aaditya et al., 2024] is openly available and a strong general biomedical baseline, but it is not adapted to oncology-specific workflows. Prior academic systems such as TrialGPT [Jin et al., 2024] approach human-level trial matching in patient-to-trial retrieval and eligibility assessment, but no fully open, end-to-end, PDAC-specific pipeline currently exists.

We argue that an *open-data, open-weight, single-workstation* pipeline is what the PDAC research community is missing. Open data lets independent groups validate and extend the work without an institutional data-use agreement; open weights permit local deployment in clinical environments where outbound data flow is restricted; and a workstation-scale recipe lowers the barrier for academic labs that do not have GPU clusters. Onca is built around these three constraints.

### Task scope

We deliberately scope the model to four PDAC workflows that are clinically high-impact and individually well-studied. *Clinical trial screening* is motivated by persistently limited pancreatic-cancer trial accrual [Guerra et al., 2021] and the fact that manual criterion-by-criterion eligibility review remains one of the dominant bottlenecks; TrialGPT [Jin et al., 2024] showed that LLMs can approach clinician performance on patient-to-trial matching, so an open-weight equivalent would let a center deploy screening on its own patient population without sending data to a third-party API. *Clinical reasoning* is motivated by PDAC’s multi-step treatment synthesis against a rapidly evolving evidence base that includes FOLFIRINOX [Conroy et al., 2011], nab-paclitaxel/gemcitabine [Von Hoff et al., 2013], and olaparib for BRCA-mutated disease [Golan et al., 2019]; existing oncology models either omit reasoning (CancerLLM is extraction-focused) or rely on private notes (Woollie), so a public-data reasoning model is needed for community evaluation of safety and grounding behavior. *Pathology report extraction* is motivated by the SEER [National Cancer Institute, 2026] and NAACCR [North American Association of Central Cancer Registries, 2016] registry requirements for structured staging, grade, margin, and lymph-node fields that are locked inside free-text reports; CancerLLM demonstrated that a fine-tuned model can recover most fields, but only on a private institution-specific corpus, so an open-data extractor aligned with the CAP Pancreas Exocrine Protocol [College of American Pathologists, 2025] is still missing. *Molecular variant evidence reasoning* is motivated by the routine use of tumor sequencing in precision oncology and the expert-curation bottleneck documented by CIViC [Griffith et al., 2017, Krysiak et al., 2023]; no open oncology LLM in our comparison set produces structured variant-to-interpretation outputs as a primary task. Together, these four tasks span the dominant LLM-amenable steps of the modern PDAC pipeline (trial enrollment, decision support, registry abstraction, molecular interpretation) and each one can be evaluated on publicly available data, which is what makes open reproduction possible.

### Contributions

This paper makes three contributions. First, we introduce an open-data, open-weight 9B PDAC model trained from publicly available sources on a four-task mixture chosen to cover the most clinically central LLM-amenable workflows in PDAC care. Second, we describe a strict train/holdout pipeline comprising 37,364 prepared training rows with per-task holdouts of 1,355 / 1,204 / 1,113 / 1,178 items, including a multi-pass-audited TCGA pathology-report annotation procedure aligned to CAP, SEER, and NAACCR conventions. Third, we report an 11-panel external evaluation against Woollie-7B, CancerLLM-7B, OpenBioLLM-8B, and the unmodified Qwopus base, with explicit per-panel metric definitions.

## 2. Methods

### Base model

We initialize from Qwopus3.5-9B-v3 [Jackrong, 2026], a dense Qwen3.5-class 9B transformer further fine-tuned (“v3”) on long chain-of-thought reasoning traces. We chose this checkpoint over alternative biomedical or general-purpose 7–9B bases for three reasons: (i) its Qwen3.5 backbone provides strong general-purpose instruction following and a tokenizer that handles long structured outputs well; (ii) its reasoning-focused post-training gives a substantially better starting point for the multi-step Clinical Reasoning task than non-reasoning bases of comparable size; and (iii) being a dense 9B model, it fits comfortably in BF16 with LoRA on a single high-memory consumer-grade GPU, avoiding the offloading overhead we observed with larger MoE candidates.

A single Unsloth BF16 LoRA adapter is attached to the frozen base; multitask behavior is communicated through prompt conditioning rather than expert routing. This keeps the released artifact compact (one adapter plus the published base) and reproducible from a single training entrypoint.

### Pipeline overview

Figure 1 summarizes the end-to-end pipeline: public source ingestion, per-task preprocessing (including the multi-pass-audited pathology annotation described in Appendix B.3), strict holdout separation, Unsloth BF16 LoRA supervised fine-tuning on the composed Qwopus3.5-9B-v3 base, and the 11-panel external evaluation.

**Figure 1:**
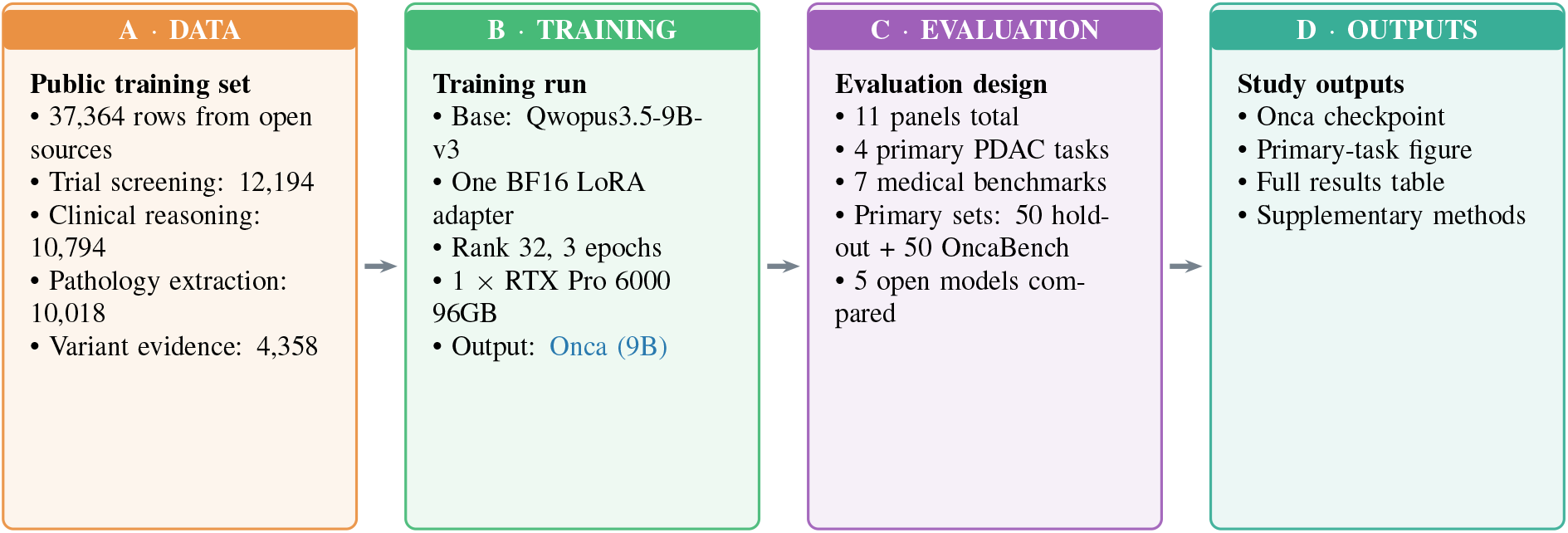
Onca pipeline. *Notes*. Overview of the Onca workflow from data preparation to evaluation. Detailed source counts are in Table 1, training settings are in Appendix A, and evaluation panels are listed in Table 4.

### Training data

The supervised run uses 37,364 examples drawn from openly available sources across the four tasks, after holdout removal. Per-task source composition is summarized in Table 1; complete data construction details, holdout protocols, and the audited pathology-extraction pipeline are documented in Appendix B.

**Table 1:** Supervised training split by task and source.

| Task | Source | Raw rows | Train |
| --- | --- | --- | --- |
| T1. Trial Screening | Kevinkrs/TrialLlama-datasets, GI/pancreatic filter [Nievas et al., 2024] | 13,549 |  |
|  | ncbi/Open-Patients, GI patient descriptions [NCBI, 2024]<sup>‡</sup> | *feeder* |  |
|  | louisbrulenaudet/clinical-trials, raw criteria [Brulenaudet, 2025]<sup>‡</sup> | *feeder* |  |
|  | *subtotal* (13,549 direct rows minus 1,355 holdout) | 13,549 | 12,194 |
| T2. Clinical Reasoning | oncollm/cancer-reasoning-traces: PDAC + capped GI non-PDAC [Hemadri et al., 2025, Jee et al., 2024] | 6,116 |  |
|  | MedReason-Stenographic [Panahi, 2025] | 3,241 |  |
|  | MedReason-GI [UCSC-VLAA, 2025] | 2,061 |  |
|  | CIViC GI evidence [Griffith et al., 2017] | 620 |  |
|  | *subtotal* (after curation & holdout removal) | 12,038 | 10,794 |
| T3. Pathology Extraction | TCGA-Reports (32 cancer types) [Kefeli et al., 2024] | 9,523 |  |
|  | TCGA GI/PAAD aligned subsets (1,433 GI + 176 PAAD) | 1,609 |  |
|  | *subtotal* (dual generic/CAP re-annotation + multi-pass audit) | 11,131 | 10,018 |
| T4. Variant Evidence | CIViC accepted evidence [Griffith et al., 2017] | 4,811 |  |
|  | CIViC submitted evidence [Krysiak et al., 2023] | 6,408 |  |
|  | MOAlmanac molecular assertions (Dana-Farber/Broad) | 556 |  |
|  | *raw pool* → 10,258 non-placeholder rows → 6,694 prompt groups → 5,536 clean candidates → −1,178 holdout | 11,775 | 4,358 |
| **Total** |  |  | **37,364** |

*Notes.* All sources are openly available. **Raw rows** are instruction-format rows before holdout removal; **Train** is the fine-tuning split after task-specific curation. Sources marked <sup>‡</sup> contribute feeder text inside T1 prompts but no standalone rows.

**Table 2:**
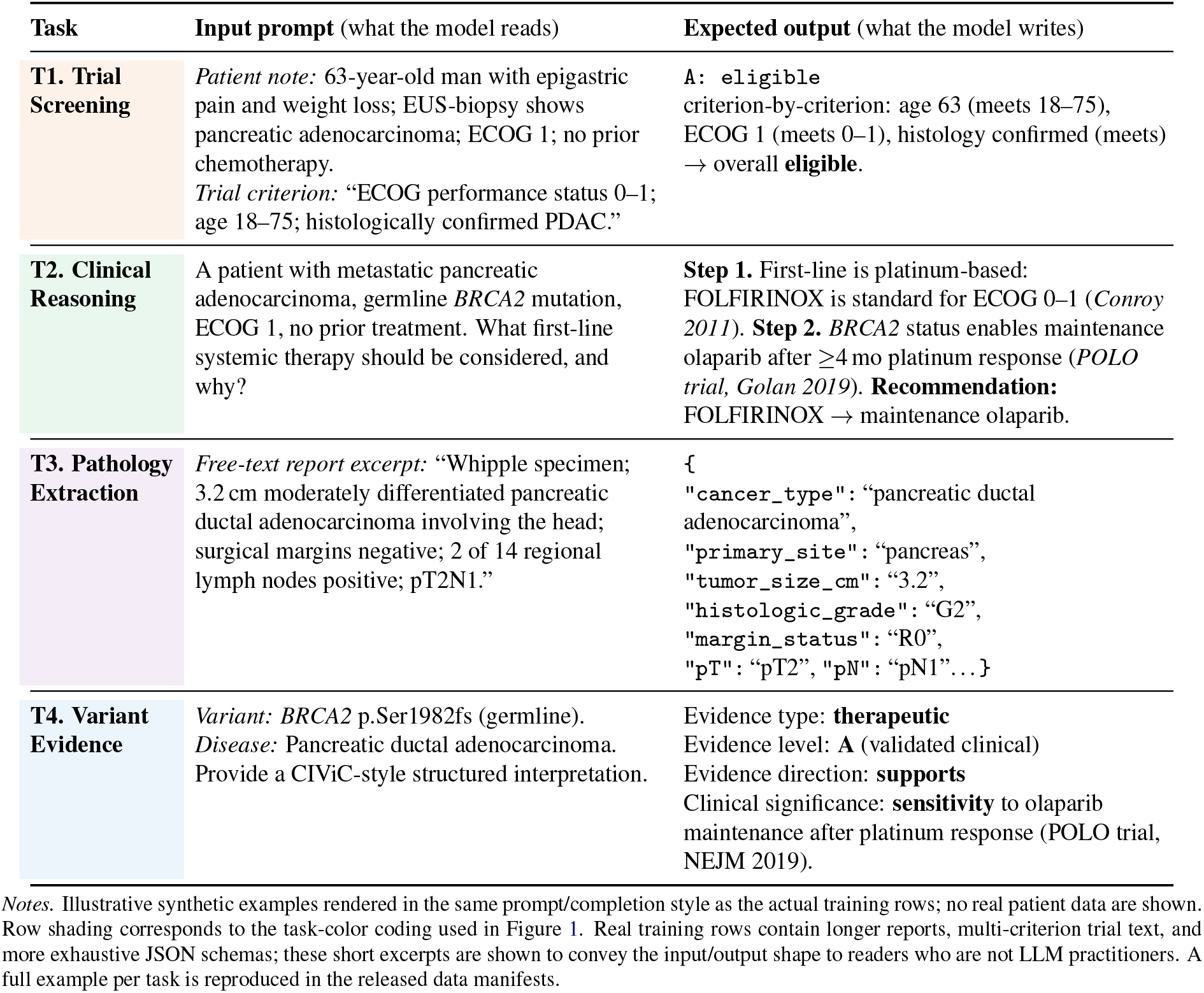
Illustrative training examples, one per task.

| Task | Input prompt (what the model reads) | Expected output (what the model writes) |
| --- | --- | --- |
| **T1. Trial Screening** | *Patient note:* 63-year-old man with epigastric pain and weight loss; EUS-biopsy shows pancreatic adenocarcinoma; ECOG 1; no prior chemotherapy.<br>*Trial criterion:* “ECOG performance status 0–1; age 18–75; histologically confirmed PDAC.” | A: **eligible**<br>criterion-by-criterion: age 63 (meets 18–75), ECOG 1 (meets 0–1), histology confirmed (meets) → overall **eligible**. |
| **T2. Clinical Reasoning** | A patient with metastatic pancreatic adenocarcinoma, germline *BRCA2* mutation, ECOG 1, no prior treatment. What first-line systemic therapy should be considered, and why? | **Step 1.** First-line is platinum-based: FOLFIRINOX is standard for ECOG 0–1 (*Conroy 2011*). **Step 2.** *BRCA2* status enables maintenance olaparib after $\geq 4$ mo platinum response (*POLO trial, Golan 2019*). **Recommendation:** FOLFIRINOX → maintenance olaparib. |
| **T3. Pathology Extraction** | *Free-text report excerpt:* “Whipple specimen; 3.2 cm moderately differentiated pancreatic ductal adenocarcinoma involving the head; surgical margins negative; 2 of 14 regional lymph nodes positive; pT2N1.” | {<br>"cancer_type": "pancreatic ductal adenocarcinoma",<br>"primary_site": "pancreas",<br>"tumor_size_cm": "3.2",<br>"histologic_grade": "G2",<br>"margin_status": "R0",<br>"pT": "pT2", "pN": "pN1"...} |
| **T4. Variant Evidence** | *Variant:* *BRCA2* p.Ser1982fs (germline).<br>*Disease:* Pancreatic ductal adenocarcinoma.<br>Provide a CIViC-style structured interpretation. | Evidence type: **therapeutic**<br>Evidence level: **A** (validated clinical)<br>Evidence direction: **supports**<br>Clinical significance: **sensitivity** to olaparib maintenance after platinum response (*POLO trial, NEJM 2019*). |

*Notes.* Illustrative synthetic examples rendered in the same prompt/completion style as the actual training rows; no real patient data are shown. Row shading corresponds to the task-color coding used in Figure 1. Real training rows contain longer reports, multi-criterion trial text, and more exhaustive JSON schemas; these short excerpts are shown to convey the input/output shape to readers who are not LLM practitioners. A full example per task is reproduced in the released data manifests.

All training sources are openly available. In Task 1, the direct rows come from TrialLlama, while Open-Patients and louisbrulenaudet/clinical-trials act as feeder corpora whose text is woven into those prompts during preprocessing rather than counted as separate examples. For all tasks, holdouts are materialized inside the same preparation pipeline that writes the train split, which keeps train and evaluation prompts separate.

The heaviest curation occurs in Task 4. Starting from 11,775 raw instruction rows, placeholder removal leaves 10,258 non-placeholder rows; grouping identical prompts yields 6,694 prompt groups, and excluding 1,158 groups with conflicting answers leaves 5,536 clean candidates before the 1,178-row holdout is applied. This is why Variant Evidence remains the smallest training stream. For Task 3, the 11,131-row English pathology mix is produced by combining TCGA-Reports with the aligned GI/PAAD subset and then applying the audited dual-prompt extraction pipeline described in Appendix B.3.

### Training procedure

We adopt the Unsloth BF16 LoRA stack with the Qwopus3.5-9B-v3 base loaded in BF16, BF16 compute, LoRA rank 32, alpha 64, max sequence length 2,048, effective batch size 64, AdamW 8-bit, cosine schedule with 5% warmup, weight decay 0.01, and learning rate 2 × 10^−4^. The run completed 3 epochs (1,752 optimizer steps), final train loss 0.7281, train runtime 47,901.0 s, and elapsed wall-clock 47,934.8 s.

### Hardware and infrastructure

Training was run on a single rented RTX Pro 6000 (96GB VRAM) instance on vast.ai. Renting a single high-memory GPU on a community marketplace — rather than maintaining an owned cluster — is a deliberate choice to demonstrate that the full pipeline is reproducible at low fixed cost. The full configuration table is reproduced in Appendix A.

### External baselines

We compare against the unmodified Qwopus base plus three external oncology/biomedical models chosen to span the design space of currently available open-weight medical LLMs: a clinically grounded oncology model (Woollie), a large-corpus extraction-oriented oncology model (CancerLLM), and a general open biomedical model (OpenBioLLM). This selection lets us separate three confounded effects — oncology-specific clinical adaptation, large institutional-scale extraction training, and broad biomedical pretraining — when interpreting our model’s behavior. Table 3 summarizes the baseline set and its core comparison axes.

**Table 3:** External baselines.

| Model | Base | Data access | Orientation |
| --- | --- | --- | --- |
| Qwopus3.5-9B-v3 [Jackrong, 2026] | Qwen3.5-9B | open | unfine-tuned internal reference. |
| Woollie-7B [Li et al., 2025a] | LLaMA-1 7B | closed (MSK) | clinically grounded 5-cancer oncology. |
| CancerLLM-7B [Li et al., 2026] | Mistral-7B | closed (UMN CDR) | 2.68M-note + 0.52M-pathology 17-cancer extraction. |
| OpenBioLLM-8B [Aaditya et al., 2024] | LLaMA-3-8B | open | general biomedical instruction tuning. |

Notes. Woollie reports AUROC 0.97 for cancer progression (MSK) with external UCSF validation and PubMedQA 0.79; CancerLLM reports F1 91.78% on its private phenotype-extraction benchmark. “Data access” refers to whether the training corpus is publicly redistributable (not just the weights).

### Evaluation panels

The evaluation is organized into two groups: four PDAC-specific primary tasks and seven medical-knowledge multiple-choice benchmarks. Each primary-task panel uses a balanced 100-item set composed of 50 holdout examples and 50 OncaBench cases. Table 4 lists the eleven panels together with their size, metric, and role; metric pairs are chosen so that each main metric is coupled to an annotation metric that exposes a common failure mode (over-verbose, schema-invalid, or parser-level failures).

**Table 4:** Evaluation panels.

| Panel | N | Main / annotation metric | Role |
| --- | --- | --- | --- |
| *Primary PDAC tasks (50 holdout + 50 OncaBench)* |  |  |  |
| Trial Screening | 50 + 50 | trial_f1 / accuracy | criterion-level eligibility judgment. |
| Clinical Reasoning | 50 + 50 | composite / key_fact_recall | case-specific treatment reasoning. |
| Pathology Extraction | 50 + 50 | overall_field_exact_match / json_parse_rate | 17-field CAP-aligned structured abstraction. |
| Variant Evidence | 50 + 50 | evidence_level_accuracy / structured_parse_rate | CIViC-style variant-to-interpretation. |
| *Secondary (cancer-/clinical-focused) MCQ benchmarks — N=50* |  |  |  |
| PubMedQA Cancer [Jin et al., 2019] | 50 | macro_f1 / accuracy | cancer-focused abstract QA. |
| MedMCQA Oncology | 50 | macro_f1 / accuracy | cancer-focused MedMCQA subset. |
| MMLU Clinical Knowledge | 50 | macro_f1 / accuracy | textbook clinical-knowledge MCQ. |
| MMLU Medical Genetics | 50 | macro_f1 / accuracy | textbook genetics MCQ. |
| *Tertiary (broad biomedical) MCQ benchmarks — N=100* |  |  |  |
| PubMedQA [Jin et al., 2019] | 100 | macro_f1 / accuracy | unrestricted biomedical QA. |
| MedQA (USMLE-style) | 100 | macro_f1 / accuracy | licensing-exam vignette reasoning. |
| MedMCQA [Pal et al., 2022] | 100 | macro_f1 / accuracy | Indian medical-entrance MCQ. |

Notes. The MCQ benchmarks are grouped into **secondary** (cancer- or clinical-focused panels at N=50) and **tertiary** (broader biomedical panels at N=100), matching the F1/F2 split used in Table 5. Clinical Reasoning composite is a 50/50 weighted median of per-row rouge\_1 and key\_fact\_recall. Variant Evidence OncaBench items are filtered to pancreatic disease and PDAC-relevant biomarkers.

## 3. Results

Figure 2 summarizes the primary PDAC evaluation. Onca leads three of the four primary task panels. On Trial Screening it reaches 81.6 F1, well above the closest baselines (Qwopus and Woollie at 50.0, CancerLLM at 45.5, OpenBioLLM at 33.3); on Clinical Reasoning it narrowly exceeds the Qwopus base (14.1 vs. 13.4) and is well ahead of the three oncology/biomedical baselines; on Pathology Extraction it slightly exceeds CancerLLM (30.5 vs. 30.0) and is well ahead of Woollie and OpenBioLLM. Variant Evidence remains the most difficult panel, where Onca reaches 6.0 and trails both Qwopus (7.2) and Woollie (25.1).

**Figure 2:**
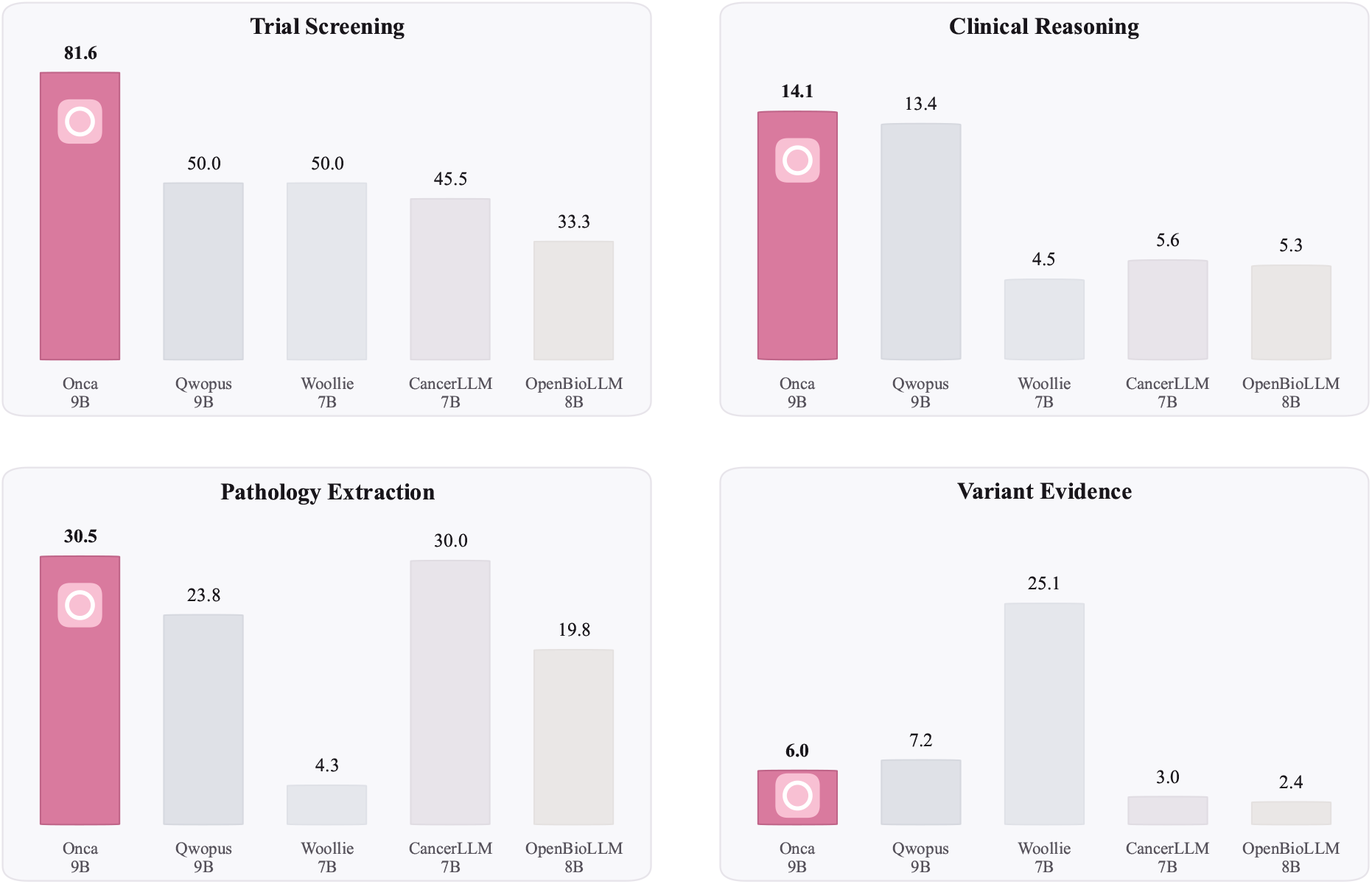
Onca four-panel primary-task comparison against external baselines. *Notes*. Four primary PDAC tasks (Trial Screening, Clinical Reasoning, Pathology Extraction, Variant Evidence Reasoning) scored on balanced 100-item sets using the task-specific metrics listed in Table 4. The seven medical-knowledge MCQ benchmarks are reported in Table 5.

Outside the primary PDAC tasks, the benchmark pattern is mixed: Onca is strongest on PubMedQA Cancer and PubMedQA, the Qwopus base remains strongest on MMLU Clinical Knowledge, and CancerLLM remains strongest on MedQA. The broader picture is selective specialization rather than across-the-board improvement. Figure 3 shows a stable supervised fine-tuning run: loss falls from 2.03 at step 10 to 0.57 by step 1,750 and flattens in the final epoch, consistent with the final reported train loss of 0.7281.

**Figure 3:**
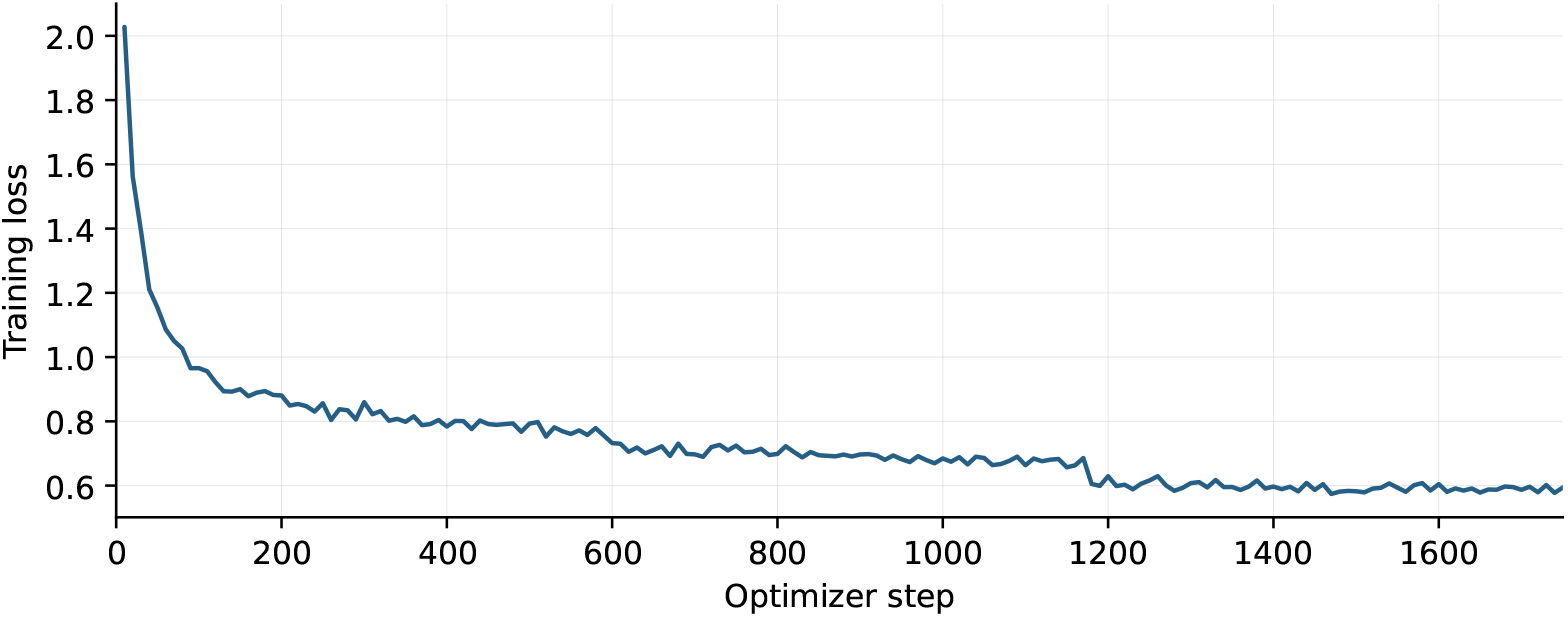
SFT training loss. *Notes*. Per-logging-step training loss for the single Unsloth BF16 LoRA supervised fine-tuning run (176 logged points, steps 10–1,750). The curve drops sharply in the first 100 steps as the adapter calibrates output formatting, then continues a slow monotonic descent over the remaining 3 epochs. The final reported train loss of 0.7281 is the trainer-reported epoch-averaged value.

Table 5 consolidates all reported metrics — the eight headline panel values, the extended per-task diagnostic metrics, the per-field pathology breakdown, and the full seven-benchmark QA matrix — into a single numerical reference so that every number cited in the text can be located in one place.

**Table 5:** Full results across primary tasks and medical-knowledge benchmarks.

| Metric | Onca | Qwopus 9B | Woollie 7B | CancerLLM 7B | OpenBioLLM 8B |
| --- | --- | --- | --- | --- | --- |
| **A. Headline panels (%-scaled; Figure 2)** |  |  |  |  |  |
| Trial Screening (F1) | **81.6** | 50.0 | 50.0 | 45.5 | 33.3 |
| Clinical Reasoning (composite) | **14.1** | 13.4 | 4.5 | 5.6 | 5.3 |
| Pathology Extraction (overall EM) | **30.5** | 23.8 | 4.3 | 30.0 | 19.8 |
| Variant Evidence (evidence-level acc.) | 6.0 | 7.2 | **25.1** | 3.0 | 2.4 |
| PubMedQA Cancer (mF1) | **68.3** | 52.2 | 38.8 | 23.9 | 52.0 |
| MMLU Clinical Knowledge (mF1) | 54.8 | **68.5** | 15.0 | 51.2 | 43.2 |
| MedQA (mF1) | 15.8 | 12.4 | 16.0 | **19.8** | 17.5 |
| PubMedQA (mF1) | **66.5** | 54.8 | 36.4 | 30.6 | 46.6 |
| **B. Trial Screening — extended (criterion-level, OncaBench 50)** |  |  |  |  |  |
| Precision | **0.909** | **0.909\*** | 0.540 | 0.532 | 0.833 |
| Recall | 0.952 | 0.667 | **1.000** | **1.000** | 0.370 |
| F1 | **0.930** | 0.800 | 0.701 | 0.694 | 0.513 |
| Macro-F1 | 0.506 | 0.055 | 0.351 | 0.253 | **0.601** |
| Eligibility accuracy | **0.660** | 0.060 | 0.540 | 0.520 | 0.620 |
| Criterion recall | **0.489** | **0.495\*** | 0.301 | 0.434 | 0.123 |
| Parse rate | 0.720 | 0.080 | **1.000** | 0.960 | **1.000** |
| **C. Clinical Reasoning — extended (per-row means, OncaBench 50)** |  |  |  |  |  |
| ROUGE-L | **0.149** | 0.147 | 0.106 | 0.121 | 0.130 |
| Key-fact recall | 0.146 | **0.157** | 0.014 | 0.033 | 0.053 |
| Treatment accuracy | **0.160** | 0.140 | 0.000 | 0.020 | 0.060 |
| **D. Variant Evidence — extended (CIViC-style schema, OncaBench 50)** |  |  |  |  |  |
| Evidence-type macro-F1 | 0.115 | **0.162** | 0.000 | 0.129 | 0.160 |
| Evidence-level accuracy | 0.120 | 0.080 | **0.240** | 0.020 | 0.020 |
| Evidence-direction accuracy | 0.300 | 0.240 | **0.640** | 0.340 | **0.640** |
| Clinical-significance macro-F1 | 0.130 | 0.063 | 0.011 | 0.034 | 0.000 |
| Summary ROUGE-L | 0.126 | **0.129** | 0.075 | 0.099 | 0.085 |
| Structured parse rate | **0.100** | 0.040 | 0.000 | **0.120** | **0.120** |
| **E. Pathology Extraction — per-CAP-field exact match (OncaBench 50)** |  |  |  |  |  |
| cancer_type | 0.08 | 0.06 | 0.02 | **0.20** | 0.18 |
| primary_site | **0.72** | 0.60 | 0.06 | 0.52 | 0.32 |

(Table 5 continued)
| Metric | Onca | Qwopus 9B | Woollie 7B | CancerLLM 7B | OpenBioLLM 8B |
| --- | --- | --- | --- | --- | --- |
| tumor_site_detail | **0.28** | 0.08 | 0.00 | 0.12 | 0.00 |
| tumor_size_cm | **0.40** | 0.38 | 0.06 | 0.26 | 0.12 |
| histologic_type | **0.46** | 0.20 | 0.06 | 0.26 | 0.18 |
| histologic_grade | **0.46** | 0.42 | 0.02 | 0.28 | 0.24 |
| margin_status | **0.54** | 0.42 | 0.08 | 0.34 | 0.28 |
| margin_distance_mm | 0.46 | **0.48** | 0.00 | 0.40 | 0.26 |
| lymphovascular_invasion | **0.58** | 0.42 | 0.10 | 0.30 | 0.26 |
| perineural_invasion | **0.52** | 0.44 | 0.06 | 0.38 | 0.26 |
| lymph_nodes_examined | **0.34** | 0.24 | 0.04 | 0.32 | 0.26 |
| lymph_nodes_positive | **0.46** | 0.38 | 0.06 | 0.36 | 0.28 |
| pT | 0.00 | 0.02 | 0.06 | **0.30** | 0.16 |
| pN | 0.00 | 0.10 | 0.02 | **0.30** | 0.18 |
| pM | 0.10 | 0.16 | 0.02 | **0.28** | 0.20 |
| ajcc_stage | 0.22 | 0.06 | 0.02 | 0.26 | **0.28** |
| additional_findings | 0.00 | 0.00 | 0.00 | 0.00 | 0.00 |
| overall_field_exact_match | **0.331** | 0.262 | 0.040 | 0.287 | 0.204 |
| json_parse_rate | 0.98 | **1.00** | 0.34 | 0.74 | 0.44 |
| field_coverage | **0.809** | 0.722 | 0.172 | 0.620 | 0.398 |
| **F1. Secondary (cancer-focused) MCQ benchmarks — N=50** |  |  |  |  |  |
| PubMedQA Cancer – acc | **0.82** | 0.68 | 0.48 | 0.36 | 0.58 |
| PubMedQA Cancer – mF1 | **0.683** | 0.522 | 0.388 | 0.239 | 0.520 |
| MedMCQA Oncology – acc | 0.08 | 0.30 | 0.18 | 0.22 | **0.34** |
| MedMCQA Oncology – mF1 | 0.138 | **0.414** | 0.216 | 0.228 | 0.392 |
| MMLU Clinical Knowledge – acc | 0.40 | **0.52** | 0.14 | 0.46 | 0.36 |
| MMLU Clinical Knowledge – mF1 | 0.548 | **0.685** | 0.150 | 0.512 | 0.432 |
| MMLU Medical Genetics – acc | 0.14 | **0.68** | 0.26 | 0.24 | 0.44 |
| MMLU Medical Genetics – mF1 | 0.239 | **0.797** | 0.324 | 0.287 | 0.516 |
| **F2. Tertiary (broad biomedical) MCQ benchmarks — N=100** |  |  |  |  |  |
| PubMedQA – acc | **0.81** | 0.57 | 0.35 | 0.30 | 0.50 |
| PubMedQA – mF1 | **0.665** | 0.548 | 0.364 | 0.306 | 0.466 |
| MedMCQA – acc | 0.19 | 0.32 | 0.12 | 0.32 | **0.34** |
| MedMCQA – mF1 | 0.304 | **0.460** | 0.178 | 0.326 | 0.399 |
| MedQA – acc | 0.09 | 0.07 | **0.13** | **0.13** | 0.11 |
| MedQA – mF1 | 0.158 | 0.124 | 0.160 | **0.198** | 0.175 |

Notes. Best value per row is shaded and bold; asterisks mark ties within rounding. Block A reproduces Figure 2. Blocks B–D report task diagnostics, Block E reports per-field pathology exact match, and Block F reports the seven medical-knowledge benchmark panels.

**Table 6:**
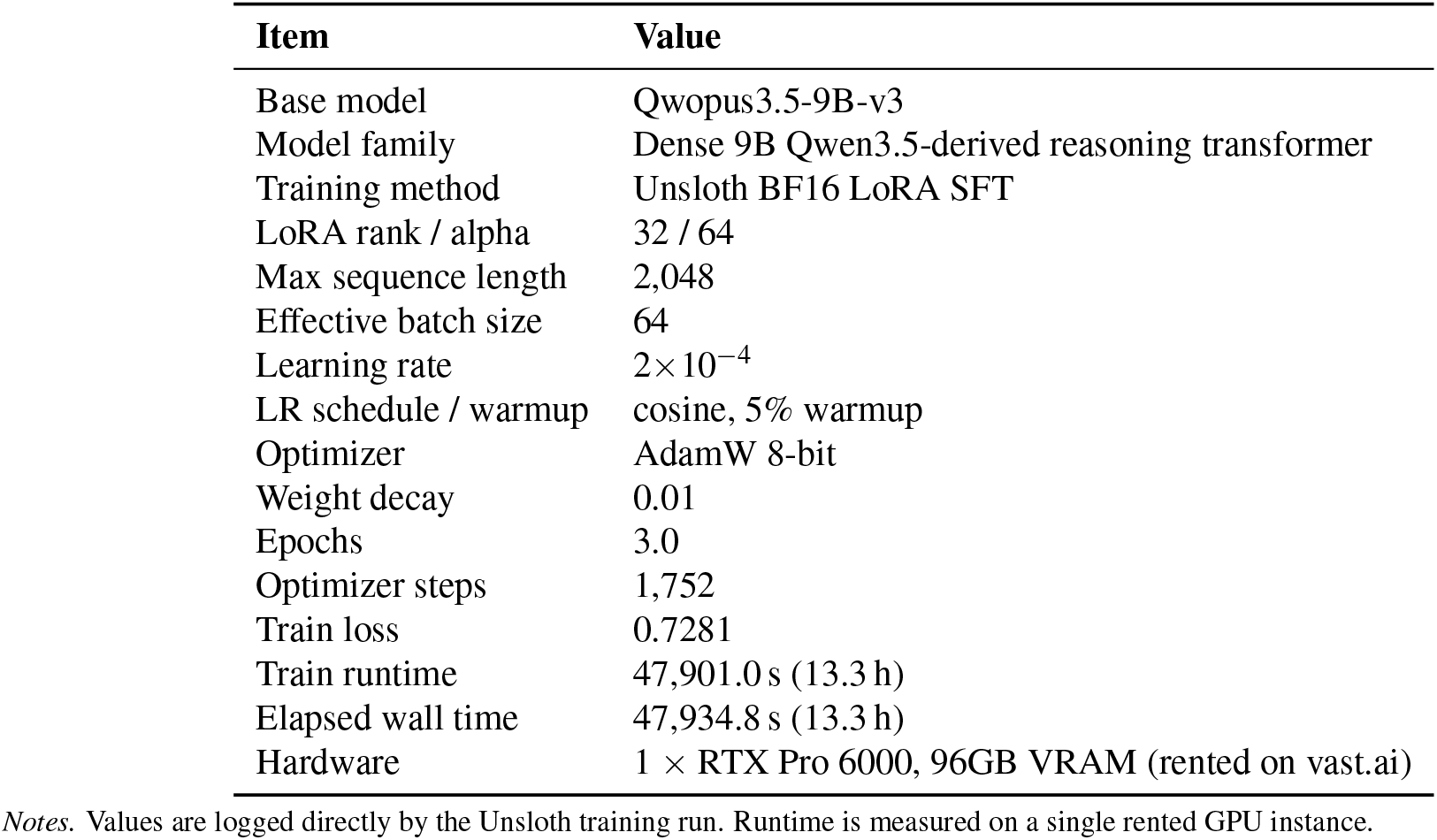
Training configuration and runtime.

| Item | Value |
| --- | --- |
| Base model | Qwopus3.5-9B-v3 |
| Model family | Dense 9B Qwen3.5-derived reasoning transformer |
| Training method | Unsloth BF16 LoRA SFT |
| LoRA rank / alpha | 32 / 64 |
| Max sequence length | 2,048 |
| Effective batch size | 64 |
| Learning rate | $2 \times 10^{-4}$ |
| LR schedule / warmup | cosine, 5% warmup |
| Optimizer | AdamW 8-bit |
| Weight decay | 0.01 |
| Epochs | 3.0 |
| Optimizer steps | 1,752 |
| Train loss | 0.7281 |
| Train runtime | 47,901.0 s (13.3 h) |
| Elapsed wall time | 47,934.8 s (13.3 h) |
| Hardware | 1 × RTX Pro 6000, 96GB VRAM (rented on vast.ai) |

Notes. Values are logged directly by the Unsloth training run. Runtime is measured on a single rented GPU instance.

## 4. Discussion

### Headline findings

The figure supports a clear but qualified conclusion: a single open 9B checkpoint, fine-tuned from public data, can lead the field on three of four PDAC-central clinical tasks and on cancer-focused biomedical QA, while remaining behind a clinically grounded oncology baseline (Woollie) on Variant Evidence and behind its own base on general clinical-knowledge MCQs. The gain on Trial Screening is large enough to be operationally meaningful but is not by itself sufficient to claim deployment readiness; criterion-level error analysis is the appropriate next step. The negative results are equally informative: Variant Evidence Reasoning is the smallest task in the training mix and uses the strictest output schema, and Woollie’s strong score on this panel suggests that institutional clinical exposure may carry signal that public CIViC/MOAlmanac evidence alone does not provide. The Qwopus base’s stronger MMLU Clinical Knowledge score is a reminder that task-focused specialization can reduce general exam-style performance, and CancerLLM’s MedQA lead suggests its broader University of Minnesota training corpus of clinical notes and pathology reports retains clinical-language coverage that our public-data mix does not match. The broader systems point is that the entire pipeline — data preparation, training, and evaluation — runs on a single rented GPU instance, which keeps independent reproduction within reach of an academic group without owned cluster infrastructure.

### Limitations

This study is intentionally narrow, and several concrete constraints shape how the results should be interpreted. First, the evaluation sample sizes are deliberately bounded: each primary PDAC panel uses 50 holdout items plus 50 OncaBench items, the four secondary MCQ panels use N=50, and the three tertiary MCQ panels use N=100. These limits reflect a deliberate decision to keep the evaluation balanced and tractable while working with relatively thin cancer-focused benchmark subsets. Because N=50 gives a 95% CI of roughly ± 14 points on an MCQ proportion around 0.5, rankings between closely performing models should be read as indicative rather than definitive. Second, the supervised fine-tuning mix uses 37,364 rows with three epochs on a single adapter, which is modest by contemporary standards and leaves the model vulnerable to mild overfitting on the more idiosyncratic training streams. This likely contributes to two observed drops: the base Qwopus checkpoint outperforms Onca on MMLU Clinical Knowledge (0.685 vs. 0.548 macro-F1), a pattern consistent with task-focused fine-tuning trading general exam-style knowledge for targeted task gains, and Variant Evidence Reasoning sits at 6.0 — its 4,358-row training pool and strict CIViC-style schema together yield more brittle outputs than the pathology or reasoning panels. Third, the model is trained on public datasets rather than institutional clinical notes, which is a strength for reproducibility but a limitation for realism: the abbreviated, context-dependent language of real-world clinical documentation [Yang et al., 2022] is only partially represented. Fourth, benchmark transfer is uneven: gains on PubMedQA Cancer and PubMedQA do not automatically translate to broader medical benchmarks. Fifth, Task 1 relies on a community reproduction (Kevinkrs/TrialLlama-datasets) rather than the official Trial-LLAMA release and exhibits a 73% irrelevant-label class skew; Task 3 has high inherent missingness in pM (∼ 47%) and AJCC stage (∼ 82%) fields, reflecting clinical reporting practice rather than annotation error; Task 4 retains the smallest train pool and should be interpreted as a focused adaptation rather than broad variant–drug coverage. Finally, Onca remains a research model — it is not validated for clinical use, does not replace clinician judgment, and should not be interpreted as a production-ready medical decision system.

## Conclusion

Onca is an open 9B PDAC-focused model trained from public data on four clinically motivated tasks. In the 11-panel evaluation it is strongest on Trial Screening, Clinical Reasoning, Pathology Extraction, PubMedQA Cancer, and PubMedQA, while remaining weaker on Variant Evidence Reasoning and not surpassing the best baseline on every benchmark. The central contribution is practical: meaningful pancreatic-cancer specialization is achievable in a fully open, single-workstation workflow.

## Data Availability

All data produced are available online

https://huggingface.co/Joesh1/onca-1.0-9B

## Ethics Statement

This study uses only publicly available, openly accessible, de-identified datasets. No data from hospitals, clinics, electronic health records, controlled-access registries, private repositories, or other non-public sources were used, and no data-use agreement was required for the corpora included here. The work does not involve intervention, recruitment, participant contact, or access to identifiable private information; it is presented as secondary analysis of non-identifiable public data only and, under standard human-subjects research definitions, does not constitute human-subjects research. The resulting model is a research artifact only and must not be used to guide clinical care without external validation, peer review, and clinician oversight.

## Acknowledgments

We are grateful to the institutions, dataset curators, and open-model developers whose publicly available resources made this work possible. We especially acknowledge the Qwen and Qwopus model developers, together with the teams who released the pathology, reasoning, trial-screening, and biomedical benchmark resources used throughout this study.

## Code and Data Availability

Model releases and related public artifacts are hosted at https://huggingface.co/Joesh1/onca-1.0-9B.

## Appendices

### A. Training Configuration

### B. Detailed Data Descriptions by Task

#### B.1 Trial Screening

Built from the Kevinkrs/TrialLlama-datasets community reproduction [Nievas et al., 2024], a 66,924-row patient–trial matching corpus in instruction format. We apply a GI/pancreatic cancer filter and retain only the train split, yielding 13,549 raw rows after deduplication, with a 73% irrelevant / 15% eligible / 12% excluded label distribution that reflects real-world screening ratios. We supplement these with ncbi/Open-Patients GI cancer descriptions [NCBI, 2024] (14,351 strict GI cancer patient descriptions) and louisbrulenaudet/clinical-trials raw criteria [Brulenaudet, 2025]. The final train split contains 12,194 rows after holdout removal. The holdout contains 1,355 items. Output is a per-criterion judgment of *eligible, excluded*, or *insufficient information*. The known limitation here is that the primary source is a community reproduction rather than the official Trial-LLAMA release, and the class imbalance can require balancing during training.

#### B.2 Clinical Reasoning

A curated 12,038-row source pool covering four supervision types:

- **oncollm/cancer-reasoning-traces** [Hemadri et al., 2025]: 3,116 PDAC reasoning traces plus a curated 3,000-example subset of GI non-PDAC reasoning traces from MSK-CHORD patients [Jee et al., 2024], generated via the OncoReason pipeline.
- **MedReason-Stenographic** [Panahi, 2025]: 3,241 compressed reasoning traces with explicit `<think>` tags (9 empty-query rows dropped).
- **MedReason-GI** [UCSC-VLAA, 2025]: 2,061 medical QA pairs with detailed reasoning, GI-filtered.
- **CIViC GI evidence** [Griffith et al., 2017]: 620 mutation–drug–disease evidence entries for GI cancers.

The final train split contains 10,794 rows after curation and holdout removal; the holdout contains 1,204 items. The 9B variant deliberately excludes weaker PubMed-abstract subsets and SFT-Mega supplements that we found degraded reasoning quality during preliminary experiments, in favor of higher-signal sources.

#### B.3 Pathology Extraction

Pathology extraction was the most labor-intensive task to prepare. The final 10,018-row train split is the output of a multi-step pipeline that we describe in detail because schema design and audit decisions materially affect the reported scores.

##### Source corpus

TCGA-Reports [Kefeli et al., 2024] provides 9,523 unannotated free-text pathology reports across 32 cancer types, of which approximately 264 are pancreatic. We additionally use 1,609 TCGA GI/PAAD aligned subsets (1,433 GI and 176 PAAD reports) where structured GDC clinical labels are available; the PAAD subset is used as ground truth for validation. One aligned record is dropped during final instruction-format materialization because it fails the downstream schema-validation pass, yielding the 11,131-row English mix used in the released 9B pipeline. The conditional NCI ML-Ready Pathology Reports collection (∼ 7,187 reports) requires NCI MoDaC portal login and is not used in the released pipeline.

##### Dual-prompt schema design

Because the source corpus is mixed-cancer rather than pancreas-only, applying a strict CAP Pancreas Exocrine schema to every report would produce systematically wrong fields for the ∼ 97% of reports that are not pancreatic. We therefore designed two complementary prompts: a *generic solid-tumor schema* that extracts 17 fields common across organ sites (cancer type, primary site, tumor site detail, tumor size, histologic type/grade, margin status/distance, LVI, PNI, lymph nodes examined/positive, pT, pN, pM, AJCC stage, additional findings), and a *pancreas-specific CAP schema* [College of American Pathologists, 2025] that constrains fields such as `primary_site to pancreas, tumor_site_detail` to a closed set (head/body/tail/uncinate/ampulla), and margin_status to the R0/R1/R2 surgical convention. A regex-based PDAC detector (matching `pancrea, PDAC, pancreatobiliary, whipple`, or `ampullary`) routes each report to the appropriate prompt.

##### Output schema and downstream alignment

The 17 structured JSON fields are aligned with the CAP Pancreas Exocrine Protocol [College of American Pathologists, 2025] for pancreas reports and remain compatible with SEER [National Cancer Institute, 2026] and NAACCR [North American Association of Central Cancer Registries, 2016] downstream registry formats. Strict per-key validation ensures every required field is present and that `cancer_type` and `primary_site` are non-empty before a record enters the train pool.

##### Validation against ground truth

Before running the full annotation pass, we validate accuracy against the 176 TCGA-PAAD aligned ground-truth pairs, comparing predicted vs. GDC values for the most error-prone fields (`ajcc_pathologic_stage, ajcc_pathologic_t, ajcc_pathologic_n`). Validation gates production runs at a per-field accuracy threshold above 70%.

##### Multi-pass audit and repair

The annotation outputs are then audited in three passes that target distinct failure modes: (i) a structural-drift audit that catches row drops, row shifts, and patient/text mismatches against source batches and high-signal semantic drift (clearly wrong margin labels, malformed staging codes against a strict regex); (ii) a repair pass that applies the corresponding corrections; and (iii) a schema/structural validation pass. The whole-dataset strict audit on the final outputs flagged 0 instances, and a fresh blind 500-row sample drew 500/500 “good” from the audit harness.

##### Known data characteristics

Even after this pipeline, a subset of staging fields remain at high “not specified” rates that reflect genuine clinical reporting practice rather than annotation error: pM is “not specified” in ∼ 47% of reports, and AJCC stage in ∼ 82%. We retain rather than impute these fields so that the trained model learns the correct “unknown” behavior.

##### Splits

The final train split is 10,018 rows after the 1,113-row holdout is excluded from the 11,131-row English mix. The holdout is the same 1,113-row set used for evaluation reporting throughout this paper, and is materialized inside the same preparation pipeline that writes the train file.

#### B.4 Variant Evidence Reasoning

The upstream source exports contain 4,823 CIViC accepted records, 6,443 CIViC submitted records [Griffith et al., 2017, Krysiak et al., 2023], and 1,014 MOAlmanac molecular assertions from Dana-Farber/Broad. After instruction-format normalization, schema harmonization, and prompt-level materialization into the released 9B SFT pool, these sources yield an 11,775-row raw Task 4 pool (4,811 CIViC accepted + 6,408 CIViC submitted + 556 MOAlmanac instruction rows). Removing placeholder variant/disease prompts leaves 10,258 non-placeholder rows. Grouping identical prompts collapses those rows into 6,694 prompt groups; 1,158 groups contain conflicting answers and are discarded, leaving 5,536 clean prompt-unique candidates, from which the 1,178-row holdout is materialized, leaving a final 4,358-row train split. This is the smallest task in the mix; claims about variant–drug coverage should remain conservative until a larger curated mapping is added.

